# Vaccine Efficacy at a Point in Time

**DOI:** 10.1101/2021.02.04.21251133

**Authors:** Dean Follmann, Michael Fay

## Abstract

Vaccine trials are generally designed to assess efficacy on clinical disease. The vaccine effect on infection, while important both as a proxy for transmission and to describe a vaccine’s total effects, requires frequent longitudinal sampling to capture all infections. Such sampling may not always be feasible. A logistically easy approach is to collect a sample to test for infection at a regularly scheduled visit. Such point or cross-sectional sampling does not permit estimation of classic vaccine effiacy on infection, as long duration infections are sampled with higher probability. Building on work by Rinta-Kokko *and others* (2009) we evaluate proxies of the vaccine effect on transmission at a point in time; the vaccine efficacy on prevalent infection and on prevalent viral load, VE_*PI*_ and VE_*PV L*_, respectively. Longer infections with higher viral loads should have more transmission potential and prevalent vaccine efficacy naturally captures this aspect. We apply a proportional hazards model for infection risk and show how these metrics can be estimated using longitudinal or cross-sectional sampling. We also introduce regression models for designs with multiple cross-sectional sampling. The methods are evaluated by simulation and a phase III vaccine trial with PCR cross-sectional sampling for subclinical infection is analyzed.

## 1. Introduction

Many vaccine trials do not directly assess vaccine efficacy on infection as there is no sampling of asymptomatic volunteers to detect presence of the pathogen. Yet the vaccine effect on infection is important to understand, both for its potential impact on transmission and to better characterize the vaccine effect on individuals. Regularly sampling trial volunteers is difficult due to the additional logistical burden imposed on volunteers and study personnel. Volunteers do periodically come in for exams and may come in to receive vaccine, blinded or unblinded, once the vaccine has been shown to be efficacious. At such visits, a sample to detect infection by e.g. a PCR test, could be collected to assess the proportion of infections in the two arms. The vaccine efficacy on the reduction in PCR positive tests at this point in time could be calculated. Yet what does it measure?

A vaccine has myriad effects and vaccine efficacy on disease, infection (susceptibility), transmission, and the population have been defined see Halloran *and others* (1999). Cross-sectional sample estimates do not accurately recover the true vaccine efficacy on infection for a person, i.e. VE_*I*_, as longer duration infections are over-represented from a singe cross-sectional sample. Rinta-Kokko *and others* (2009) introduced the vaccine efficacy on pneumococcal carriage which we call the vaccine efficacy on prevalent infection or VE_*PI*_. This should be a better proxy for the effect of vaccine on transmission than VE_*I*_, as it reflects that longer infections have more transmission potential. We also introduce the vaccine efficacy on prevalent viral load or VE_*PV L*_. This is the proportion reduction in the amount of virus in a vaccinated person at a point in time. Since more virus should increase transmissibility, this may be an even better proxy for transmission.

In this work, we motivate and provide simple estimates of these three aspects of vaccine efficacy. We apply a proportional hazards model for the instantaneous risk of infection over time and show how VE_*I*_, VE_*PI*_ and VE_*PV L*_ are expressible as functions of the parameters of this model which is a form of a mark specific hazard, see Gilbert *and others* (2004), and has been previously used to estimate vaccine efficacy on the number of founding viruses of an infection, see Follmann and Huang (2015). The model allows us to demonstrate how VE_*PI*_ and VE_*PV L*_ can be estimated with longitudinal e.g. biweekly sampling and can be estimated using cross-sectional data, provided the vaccine to placebo ratio of mean infection duration, or viral load is known.

This generalizes results connecting between incidence, duration, and prevalence to viral loads see Freeman and Hutchison (1980), Keiding (1991). We show how to combine multiple cross-sectional samples by formulating regression models for infection and viral load given infection. These models readily incorporate covariates such as baseline variables and functions of time since vaccination and provide adjusted estimates of VE_*PI*_ and VE_*PV L*_. We evaluate the estimators via simulation and analyze the day 28 cross-sectional sample data from the phase III Moderna COVID-19 vaccine trial Moderna (2020).

## 2. Motivation

Vaccine efficacy for infection (susceptibility) typically requires frequent evaluation. For example, for COVID-19 vaccine trials twice weekly PCR testing might be required to ensure nearly all infection events are captured. For large studies with a rare disease, a good estimate of the vaccine efficacy on infection is

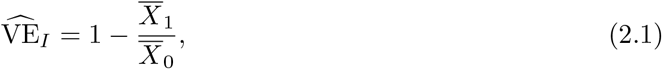

where 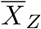 is the observed proportion of infections recorded in the volunteers from arm *Z* measured over a common period of time. In practice this might be all volunteers, baseline seronegative volunteers, or baseline seronegative volunteers without prior symptomatic disease, depending on the question.

Frequent sampling for infection can be burdensome and expensive. A logistically easy approach is to sample subjects at a single point in time such as a crossover or serology visit. Define 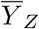 as the proportion of tested individuals in arm *Z* with an infection at such a visit. We can form a simple estimate of vaccine efficacy for infection at this point in time as

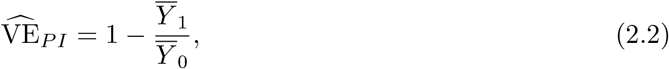

which we call the vaccine efficacy for prevalent infection or VE_*PI*_. This approach was introduced by Rinta-Kokko *and others* (2009) to describe the vaccine effect on pneumococcal carriage, though their metric was the odds ratio, also see Thompson *and others* (1998). This differs from 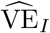. If the vaccine tends to make the duration of infection shorter, then fewer vaccine infections will be collected when sampled at a single point in time as shown in Figure 1. As suggested by Rinta-Kokko *and others* (2009), 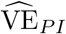 may be a better measure of transmission risk to the community as it reflects the reduction in the number of infected individuals in the community on a given day. Suppose the vaccine had no effect on infection but reduced duration of infection by 90%. Then on any given day, there would be 90% fewer vaccine volunteers who were PCR positive with presumably less risk of transmission into the community. In contrast, 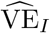 measures the effect of vaccination on an individual’s risk of infection (regardless of duration).

**Fig. 1.**
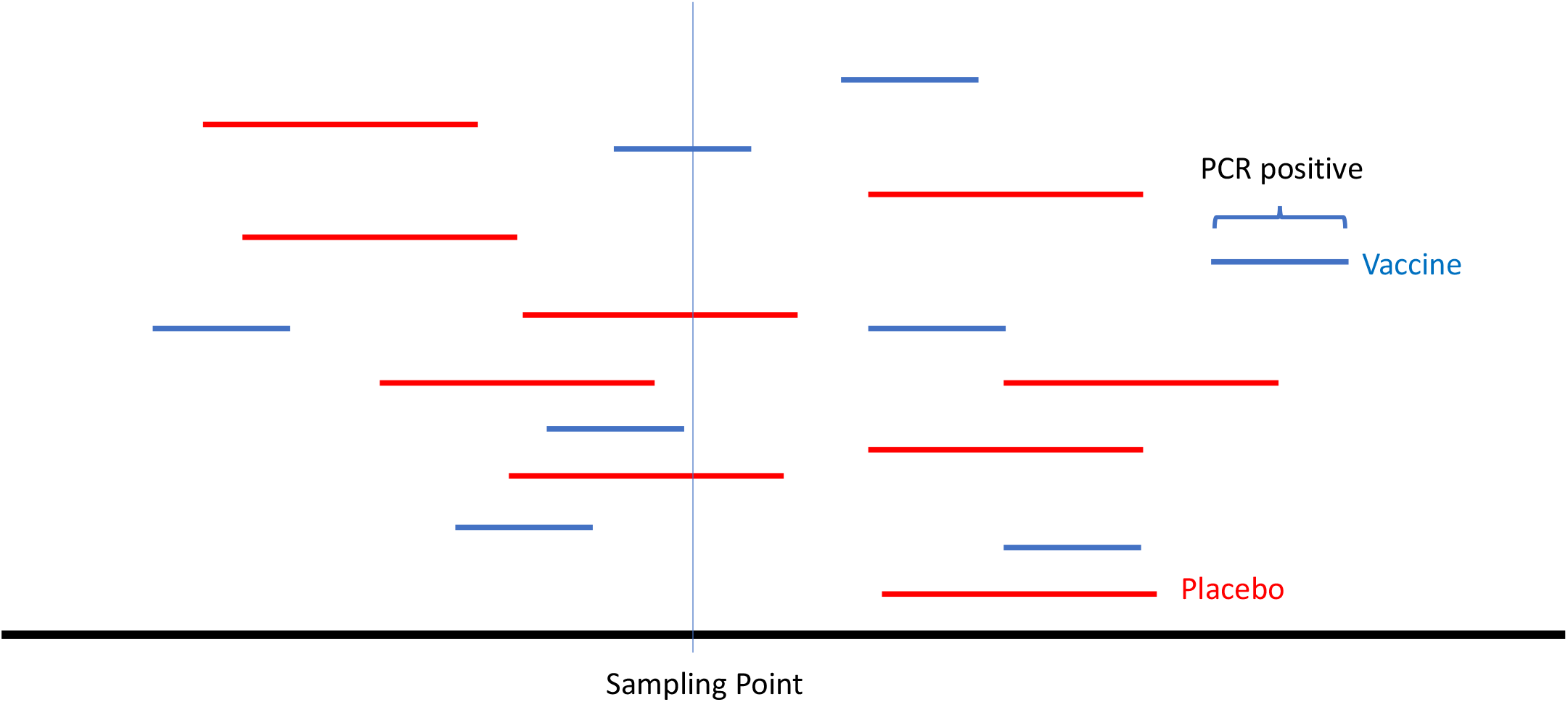
Two different estimates of vaccine efficacy for infection. The vaccine efficacy for prevalent infection or 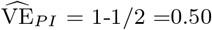. The traditional vaccine efficacy counts all infections and is 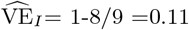.

If the cross-sectional sample includes a measure of viral load we can create an even better proxy for the vaccine effect on transmission. To motivate this estimate, suppose the vaccine had no effect on infection, nor duration, but reduced the viral load during infection by 90%, compared to placebo. Then on any given day, there would be 90% less virus in the vaccine volunteers compared to placebo volunteers. Presumably this would translate into a substantially reduced risk of onward transmission. We thus form a simple estimate of the vaccine efficacy on viral load as the proportion reduction in total viral load at a point in time of vaccine effiacy on prevalent viral load as

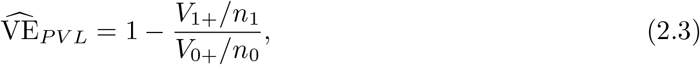

where *V*_*Z*+_ is the sum of viral loads over all sampled volunteers on arm *Z* and *n*_*Z*_ the number sampled. Let *Y*_*Z*+_ be the number of infections on arm *Z* from the cross-sectional sample. Because 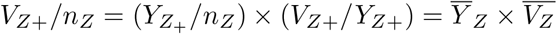, (2.3) can be written as

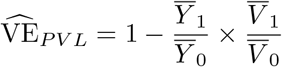

Thus 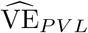 blends a vaccine effect on the probability a person is infected on a given day times an effect on the mean viral load among the infecteds.

In the next section we develop an infection process model and formally define VE_*PI*_, VE_*PVL*_, and VE_*I*_ in terms of the parameters of this single process. This representation demonstrates how to estimate these metrics from a longitudinal study and also how VE_*I*_ can be estimated from a single cross-sectional study—provided we have some auxiliary information.

## 3. Theoretical Development

Let *T* be the time from vaccination to the start of infection. We assume a proportional hazards model where the hazard for infection is given by

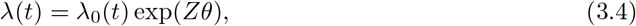

where *Z* is the vaccine indicator, *t* the time since vacccination, and *λ*_0_(*t*) an unspecified basleine hazard. Let *S*(*t*) = *P* (*T > t*) be the survivor function.

The hazard function representation can be decomposed as *λ*_0_(*t*) = *ω*(*t*)*P* (*A* = 1|*Z* = 0), and *θ* = log{*P* (*A* = 1|*Z* = 1)*/P* (*A* = 1|*Z* = 0)} where *A* is the indicator of acquisition of infection *given* exposure, and *ω*(*t*) is the exposure process common to both groups, as shown in Follmann and Huang (2015). Thus 1 *−* exp(*θ*) = 1 *− P* (*A* = 1|*Z* = 1)*/P* (*A* = 1|*Z* = 0), and we define vaccine efficacy against infection as VE_*I*_ = 1 *−* exp(*θ*). With frequent longitudinal assessments of infection, we can estimate VE_*I*_ by maximizing the partial likelihood as is traditional for the Cox regression model. A simpler estimate of exp(*θ*) is to directly use (2.1) which, in a large trial with a rare disease, approximates the simple hazard ratio estimate described in Machin and Gardner (1988).

We next use this model to define VE_*PI*_ and VE_*PV L*_ parameters. The probability of an infection starting during a small interval of length (say a day) for a randomly selected volunteer is approximately *λ*(*s*)*S*(*s*) and for a randomly selected uninfected volunteer at day *s* is *λ*(*s*)*ϵ*. For a rare disease *S*(*s*) is approximately 1 so whether we condition on infection or not does not matter.

The probability of an active infection at a given point in time *s* requires that the infection occur prior to *s* and be detectable at time *s*. To derive this probability, suppose placebo infections have duration at most 3 days with probabilities *p*_0_(1), *p*_0_(2), *p*_0_(3) respectively, and that *λ*_0_(*s*) = *λ*_0_(*s −* 1) = *λ*_0_(*s −* 2). To simplify, suppose that infections occur at the start of the day. If a person infected at day *s −* 2 has a duration of 3 days, then they will be tallied as an infection at day *s*. Persons infected on day *s −* 1, with durations of 2 or 3 days will be detected on day *s*, and everyone infected on day *s* will be detected. Thus for large studies with a rare disease and small *λ*_0_(*s*), the probability an infection in the placebo arm is detected on day *s* is approximately *λ*_0_(*s*)[*p*(3) + {*p*(3) + *p*(2)} + {*p*_0_(3) + *p*_0_(2) + *p*_0_(1)}] which equals 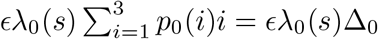, where by definition, Δ_0_ is the mean placebo duration. This argument generalizes beyond 3 days and applies to the vaccine group as well. Thus the probability of an active infection at day *s* in arm *Z* is approximately

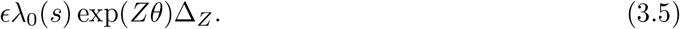

Given (3.5) we can deduce that the ratio of the probability of an active infection from someone on vaccine divided by the probability of an active infection from someone on placebo at time *s* is approximately

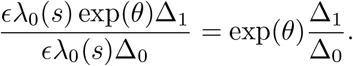

This well known result relates prevalence to incidence and duration see e.g. Freeman and Hutchison (1980), Keiding (1991). We define the vaccine efficacy for prevalent infection as

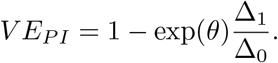

With a longitudinal study with frequent sampling we identify the time of the start of each infection. With frequent post-infection sampling we can also record the duration of each infection. This allows estimation of *θ* using Cox regression, and estimation of Δ_0_ and Δ_1_ using the sample mean durations which thus allows an estimate of VE_*PI*_. So a conventional longitudinal study designed to estimated VE_*I*_ can also report an estimate of VE_*PI*_, the putatitvely better proxy for transmission.

We can also directly estimate VE_*PI*_ from the cross-sectional study using (2.2) as 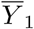 and 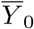 in (2.2) provide unbiased estimates of *λ*_0_(*s*) exp(*θ*)Δ_1_ and *λ*_0_(*s*)Δ_0_, respectively. Since VE_*P I*_ is free of *s*, the sampling time for different individuals can be chosen to be logistically convenient.

We next define VE_*PV L*_. Let *V* be the viral load at the time of sampling. We can think of *V* as being selected via a two stage process. Imagine the set of all infections. First we select an infection with probability proportional to *D*, the duration of the infection. Then we randomly pick a day and record the viral load. As before, suppose the placebo group has durations *D* = 1, 2, 3 with probabilities *p*_0_(1), *p*_0_(2), *p*_0_(3) and let *E*{*V*_0_(*i*)|*D*} be the mean viral load on day *i* = 1, …, *D* for infected placebo volunteers who have *D* days of detectable viral load. A placebo person infected on day *s−*2 has their infection detected on day *s* if they have *D* = 3 which occurs with probability *p*_0_(3) and such people have have mean viral load on day *s* of *E*{*V*_0_(3)|*D* = 3}. Building on this reasoning and the arguments used to derive Δ_*Z*_, one can show that the expected viral load in over the entire the placebo group at the time of the cross-sectional sampling is approximately

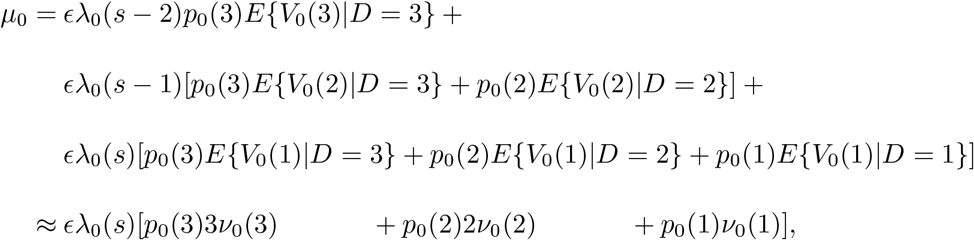

where 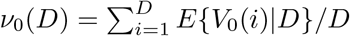 is the mean viral load over the *D* days for placebo volunteers with durations of length *D*. This argument generalizes and we can represent *µ*_*Z*_ as

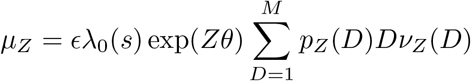

where *M* is the maximum duration. We thus deduce that at time *s* the expected amount of virus from a vaccine volunteer (whether infected or not) divided by the expected amount of virus from a placebo volunteer (whether infected or not) is

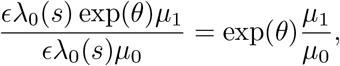

and we define the vaccine efficacy for prevalent viral load as

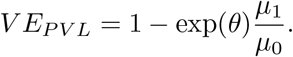

We note that if the average viral load is independent of duration so that *ν*_*Z*_(*D*) = *ν*_*Z*_ we obtain the nice expression

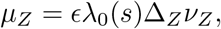

which is simple, but may not hold in practice, as e.g. volunteers with longer durations may have higher peak viral loads and possibly higher *ν*_*Z*_(*D*).

As for VE_*P I*_, with a longitudinal study we can obtain estimates of *θ* using Cox regression and with daily post infection sampling of viral loads, obtain estimates of *µ*_0_, *µ*_1_. A simple cross-sectional sample estimate of VE_*PV L*_ is given by (2.3) as *V*_1+_*/n*_1_ and *V*_0+_*/n*_0_ provide unbiased estimates of *λ*_0_(*s*) exp(*θ*)*µ*_1_ and *λ*_0_(*s*)*µ*_0_, respectively.

With this machinery, other proxies for transmission can be formed. For example, if transmission were unlikely to occur unless the viral load exceeded a threshold, say 5 logs, then we could redefine viral load as *V* ^*\**^= *I*(*V >* 5) where *I*() is the indicator function. Or if there were a known function say *P* (*V*) that provided the probability of transmission given a viral load of *V*, we could use the function *P* (*V*) instead of *V* in the above development.

Inference for Cox based longitudinal estimates of VE_*PI*_ and VE_*PV L*_ follows from the approach discussed in Follmann and Huang (2015). That work blended a Cox estimate on the hazard ratio for infection with the ratio of the mean number of founding viruses given infection, thus defining a vaccine efficacy on the mean number of founder viruses. The delta-method was proposed for approximate inference which applies to both VE_*P L*_ and VE_*PV L*_. In addition, a weighted estimatsing equations approach for the integer virus counts was proposed which could be directly used for VE_*PI*_ as the durations are integers. We assume the longitudinal study also captures all incident infections with daily sampling.

To form confidence intervals for the simple crosssetional estimates of VE_*PI*_ and VE_*PV L*_ given by (2.2) and (2.3), respectively one could use the delta-method or the bootstrap, or if necessary, methods crafted for small samples such as a non-informative Bayesian approach.

While a pure cross-sectional study cannot estimate Δ_*Z*_, if the ratio Δ_1_*/*Δ_0_ were known or estimated from a different study, one could craft an estimate of the classic metric VE_*I*_ that ignores infection duration as

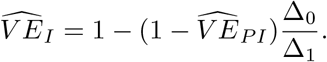

If there were no data to directly estimate Δ_1_*/*Δ_0_, one might specify different plausible values of Δ_1_*/*Δ_0_ to obtain a range of VE_*PI*_ estimates.

A concern with COVID-19 vaccines is that they might tend to shift symptomatic infections to asymptomatic infections which might increase transmission potential as silently infected individuals might not socially distance Mehrotra *and others* (2020). To examine this possibility, assume that a proportional hazards models holds for the risk of acquisition of disease as in (3.4), but with parameter *θ*_*D*_. For clarity, denote the *θ* of (3.4) as *θ*_*I*_ Now if the vaccine has the same effect on asymptomatic and symptomatic infections then *θ*_*I*_ = *θ*_*D*_. To test *H*_0_ : *θ*_*I*_ = *θ*_*D*_, we can estimate 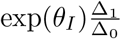 using the cross-sectional sample and estimate exp(*θ*_*D*_) directly from continuous monitoring for disease from the same trial. If the ratio Δ_1_*/*Δ_0_ were known or estimable, we could fashion a test of *H*_0_ : *θ*_*I*_ = *θ*_*D*_ by forming a Wald statisic

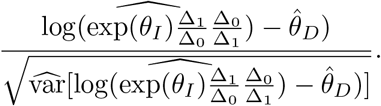

As before, the delta-method or bootstrap could be used to estimate the variance in the denominator and the Wald statistic compared to a standard normal null distribution.

## 4. Regression And Multiple Cross-sectional Samples

The above development was for a single cross-sectional sample without covariates other than the vaccine indicator. To accommodate multiple samples while allowing for additional covariates leads naturally to regression modeling for correlated data.

Let *Y*_*Zki*_ be the indicator that person *i* at visit *k* in arm *Z* is positive. A binomial model with a log-link can be used to model the probability of infection. This model, as opposed to a logit model, has the advantage that it readily allows an estimate of vaccine efficacy as one minus a relative risk. As an example, a flexible specification that can allow for waning efficacy is given as

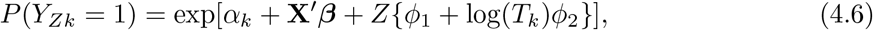

where *T*_*k*_ is the actual time post vaccination of visit *k* for a given subject, **X** are baseline covariates, *α*_*k*_ a visit specific offset, and *φ*_1_, *φ*_2_ specify a log-linear form of waning efficacy. Since volunteers are sampled over multiple visits, the outcomes from an individual might be correlated so one can fit generalized estimating equations (GEE) model with the individual as a cluster, see Zeger and Liang (1986). The vaccine efficacy on prevalent infection at visit *k* from (4.6) is given by

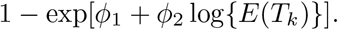

where *E*(*T*_*k*_) is the average time since vaccination among those who are sampled at visit *k*.

One can always perform these analyses within subsets of subjects or define the outcome to be different from being PCR positive. For example, we might apply (4.6) to seronegative subjects at each point in time, or to subjects who have had no prior evidence of any infection. Previous infection could be documented by serology, previous subclinical infection, or previous COVID-19 disease. If it were known that only a single infection were possible, one could eliminate previously infected individuals from the analysis, rather like eliminating cases from the risk set in the partial likelihood of Cox regression.

For multiple cross-sectional viral loads the development is slightly more complex. One way is to develop models conditional on infection, rather like a hurdle model, see Cragg (1971). Analogous to (4.6), we might specify a log-linear model as

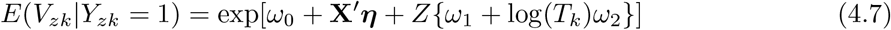

If an individual were positive at multiple visits a GEE approach could be used with individual as the cluster.

The regression based VE_*PV L*_ at visit *k* is given by

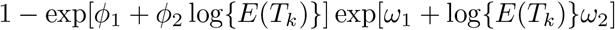

Under our model, the parameter estimates from (4.6) and (4.7) are independent. Thus the delta-method could be used for testing and construction of confidence intervals. A simpler approach would be to bootstrap individuals. The use of a log-linear specification for (4.7) was motivated by the nice cancellation of the *ω*_0_ and *η* parameters in the regression based VE_*PV L*_ parameter. In practice, a linear or other type of model may fit the data better resulting in a different form for VE_*PVL*_.

## 5. Simulations

In this section we evaluate the cross-sectional sample estimates under a few different scenarios meant to roughly approximate COVID-19 vaccine trials. A large clinical trial was assumed with 15,000 per arm and a total of 500 infection times were generated uniformly over the interval 0 to 200 with VE_*I*_ set at 0.00, 0.50, or 0.75. We assume that the viral load trajectory of an infected placebo volunteer follows a two stage model, with mean trajectory rising to 6 logs over the course of 4 days followed by a linear decline to zero over a mean of 28 days, see To *and others* (2020). Vaccine infections were similar but with a peak viral load of 6 or 4 logs and a mean time to zero of 14 or 28 days. Each infected volunteer drew random deviations from the mean linear rise and mean time to zero which were Gaussian with standard deviations 0.125, 2.00, respectively with correlation 0.50. Each day the measured viral load for an infected individual was given by the mean viral load plus Gaussian error with a standard deviation of 0.10. Infection ended when the simulated viral load was first less than zero. We approximated the true Δ_0_, Δ_1_, *µ*_0_, and *µ*_1_ by simulation of 100,000 infection episodes per arm which allowed us to approximate the true VE_*PI*_ and VE_*PV L*_.

A cross-sectional study was conducted on day 100. Any volunteer with a detectable viral load on day 100 was tallied and their viral load recorded. Table 1 presents the results. The estimates are close to the true values. The average 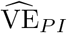 increases as the vaccine induced duration decreases and the average 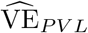 increases as the vaccine induced peak decrease. The placebo mean viral load 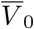 is the same over all scenarios—as the placebo generation model is the same. The vaccine mean viral load 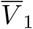 decreases with a lower peak viral load, but stays the same if the peak is unchanged but the duration is changed. The variances of 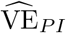 and 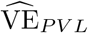 decrease with increasing VE. In contrast, the variance of 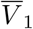 increases with increasing VE as there are fewer events with which to estimate *µ*_*Z*_. For fixed VE_*I*_, the variance of 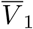 is smallest for the scenarios with a peak viral load of 4 and duration of 28 days.

**Table 1.** Simulated performance of the simple cross-sectional estimates of VE_*PI*_ and VE_*PV L*_. The mean number of infections per arm is recorded along with the mean VEs and viral loads. Each row is based on 1,000 simulated trials. The placebo arm has a mean peak viral load of 6 and a mean duration of 28 days for all scenarios. Each pair of rows reports the Monte Carlo means and (variances).

| Inputs |  |  |  |  | Performance |  |  |  |  |  |
| --- | --- | --- | --- | --- | --- | --- | --- | --- | --- | --- |
| $VE_I$ | $VE_{PI}$ | $VE_{PVL}$ | Peak | Infection | # Positive | | $\widehat{VE}_{PI}$ | $\widehat{VE}_{PVL}$ | $\bar{V}_0$ | $\bar{V}_1$ |
|  |  |  | VL | Duration | Vaccine | Placebo |  |  |  |  |
| 0.00 | 0.00 | 0.00 | 6 | 28 | 34.30<br>(32.1) | 33.90<br>(35.5) | -0.047<br>(0.071) | -0.067<br>(0.101) | 3.04<br>(0.087) | 3.07<br>(0.098) |
| 0.50 | 0.75 | 0.75 | 6 | 14 | 11.10<br>(11.5) | 45.50<br>(39.7) | 0.750<br>(0.007) | 0.740<br>(0.011) | 3.04<br>(0.066) | 3.13<br>(0.298) |
| 0.50 | 0.50 | 0.67 | 4 | 28 | 22.70<br>(22.4) | 45.50<br>(39.7) | 0.490<br>(0.018) | 0.654<br>(0.011) | 3.04<br>(0.066) | 2.04<br>(0.067) |
| 0.50 | 0.75 | 0.83 | 4 | 14 | 11.10<br>(11.5) | 45.50<br>(39.7) | 0.750<br>(0.007) | 0.826<br>(0.005) | 3.04<br>(0.066) | 2.09<br>(0.137) |
| 0.75 | 0.88 | 0.88 | 6 | 14 | 6.74<br>( 6.5) | 54.50<br>(50.9) | 0.874<br>(0.002) | 0.868<br>(0.003) | 3.04<br>(0.059) | 3.15<br>(0.548) |
| 0.75 | 0.75 | 0.83 | 4 | 28 | 13.80<br>(12.7) | 54.50<br>(50.9) | 0.742<br>(0.006) | 0.825<br>(0.003) | 3.04<br>(0.059) | 2.05<br>(0.111) |
| 0.75 | 0.88 | 0.92 | 4 | 14 | 6.70<br>( 6.5) | 54.50<br>(50.9) | 0.874<br>(0.002) | 0.912<br>(0.001) | 3.04<br>(0.059) | 2.11<br>(0.255) |

We selected the last simulated dataset from the last row of Table 1 to illustrate an analysis. Figure 3 provides the generated viral load trajectories of the infected volunteers and projects the day 100 viral load onto the right axis. For this setting, the true VE_*I*_ =0.75 and the vaccine reduces peak viral load by 2 logs and cuts the duration in half. There were a total of 6 infections on vaccine and 43 on placebo for a cross-sectional estimate of 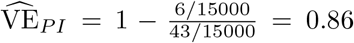. The mean viral loads on the two arms were 2.06 and 3.07. Thus 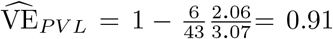. The variances of the vaccine and placebo viral loads were 0.33 and 4.47 respectively, though the standard errors of the mean were more similar at 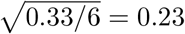 and 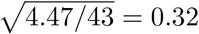. We use the percentile bootstrap to form confidence intervals. With 10,000 bootstrap samples the 95% confidence intervals are (0.71,0.96) and (0.80,0.95), respectively, for VE_*P I*_ and VE_*PVL*_.

**Fig. 2.**
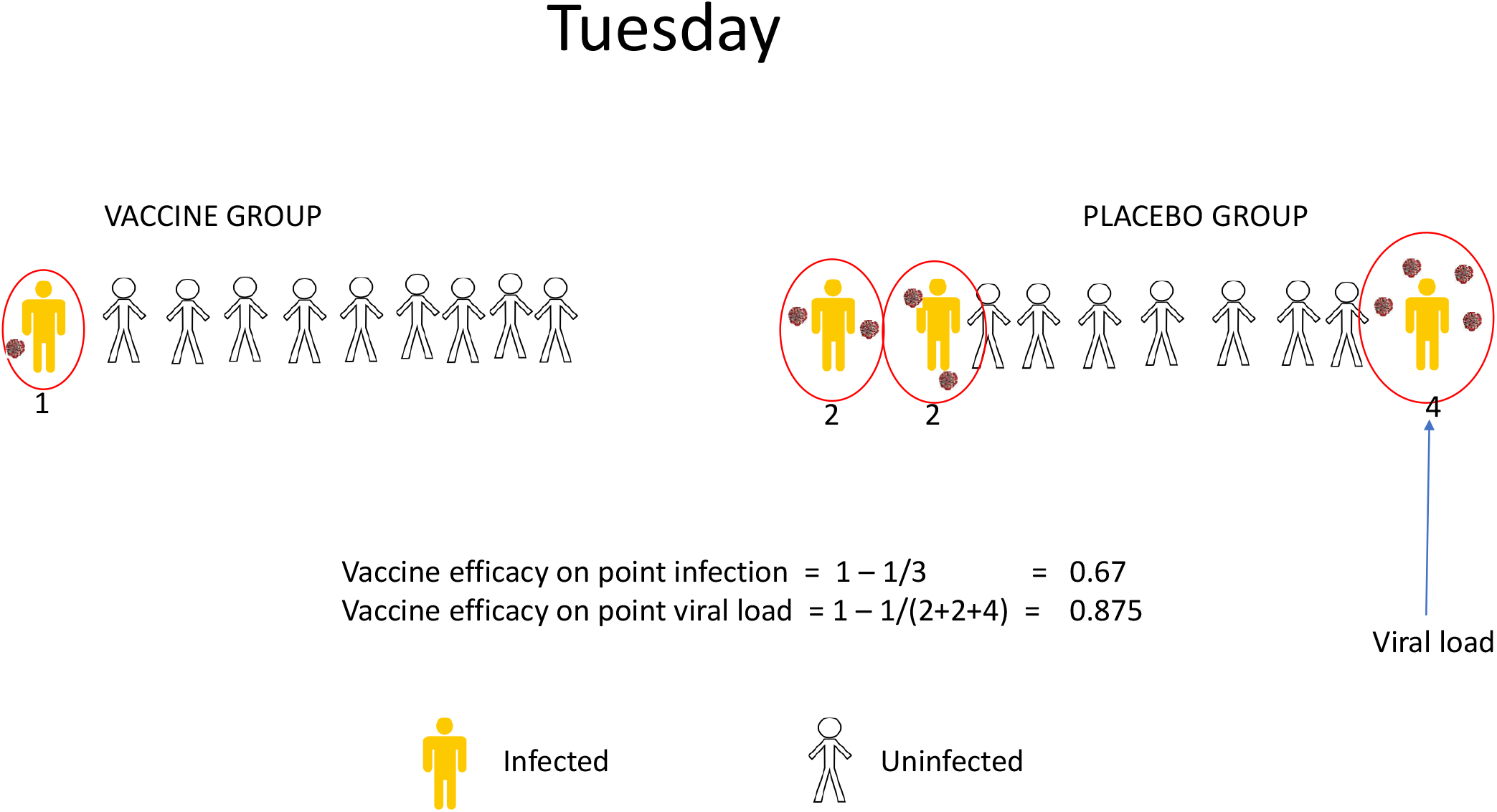
Vaccine efficacy estimates at a point in time. The placebo group has more infected and they have higher viral loads. The placebo group should be more infectious both when we contrast total infecteds and especially when we contrast total viral load. Two metrics reflect this idea. The vaccine efficacy for prevalent infection is estimated as 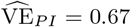. The vaccine efficacy for prevalent viral load is estimated as 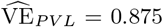.

**Fig. 3.**
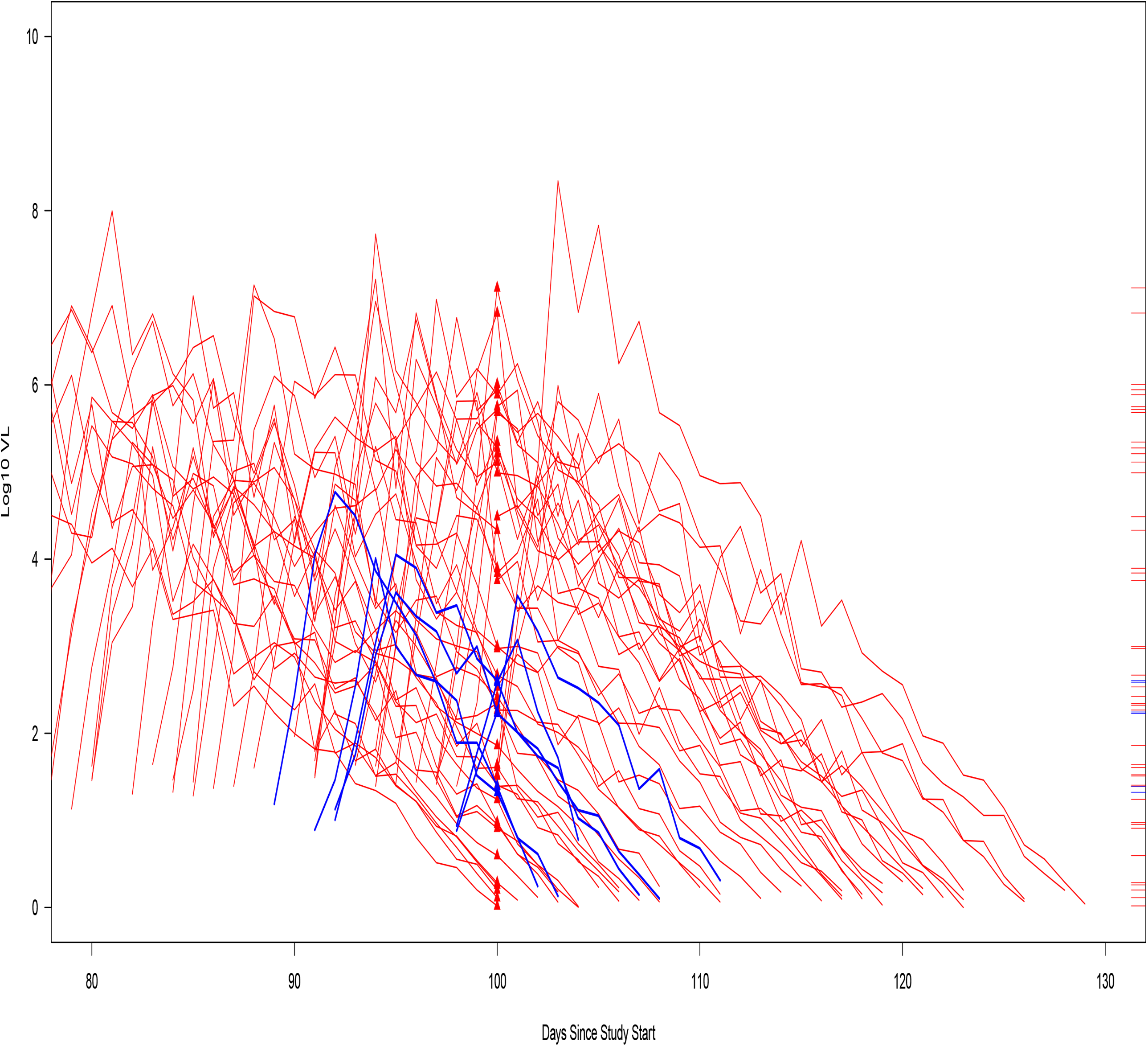
Simulated data set for a cross-sectional study with true VE_*I*_ =0.75, where vaccine reduces the peak viral load by 2 logs and the duration of infection by 14 days. Samples are drawn at day 100 to identify infections and viral loads, thus only the tics on the far right are observed. Blue denotes vaccine and red placebo.

In the supplementary materials, we examine the sensitivity of the simulation results to the assumption of a constant attack rate before the day of point samp. Given our durations are at most about 30 days, we varied the attack rate to either increase 3 fold or reduce by a factor of 3 from day 70 to day 100. Based on the pandemic in the United States, a factor of 3 seemed a plausible worst case scenario. The impact on estimates of VE_*P I*_ and VE_*P V L*_ was modest with less than a 10% bias under the least favorable setting. The mean viral loads increased about 15% under the increasing attack rate setting and decreased about 10% under the decreasing attack rate setting.

## 6. Example

The Cove trial randomized approximately 30,000 volunteers equally to two doses of the vaccine mRNA-1273 or placebo delivered 28 days apart. The trial was designed to achieve 150 cases of symptomatic COVID-19 disease which was actively monitored, see Moderna (2020) At the second vaccination visit, serology and PCR sampling were performed to identify subjects who were both serologically and PCR negative. The trial reported high efficacy on the primary endpoint of symptomatic disease at the first interim analysis and quickly applied for an Emergency Use Authorization from the Food and Drug Administration. Among baseline seronegative volunteers, the reported PCR positive rates at day 28 for asymptomatic volunteers, were 14/14134 and 38/14073, respectively for the vaccine and placebo arms, see moderna (2020). Using (2.2), we estimate VE_*PI*_ as 0.63. A 95% bootstrap confidence interval based on 10,000 bootstrap samples is (0.35, 0.82). Thus using a cross-sectional design provides useful information about the efficacy of 1 dose of the mRNA-1273 vaccine on silent infections that do not progress to symptomatic disease.

While the mean duration of silent infections is not known for unvaccinated nor mRNA-1273 vaccinated volunteers, we can specify some ratios Δ_1_*/*Δ_0_ to give a range of plausible estimates of the traditional VE_*I*_ metric. We illustrate by postulating that Δ_1_*/*Δ_0_ ranges from 0.50 to 1.00. That is, the vaccine cuts the mean duration in half, leaves it unchanged, and everything in between. This results in associated VE_*I*_ estimates ranging from 1 *−* (14*/*14134)*/*(38*/*14073) *×* 2 = 0.27 to 0.63.

## 7. Discussion

Necessity can be the mother of invention. This work was motivated by the necessity of interpreting infection counts and viral loads from a single cross-sectional sample of asymptomatic trial volunteers. We demonstrate that such samples can be used to estimate two meaningful metrics; the vaccine efficacy on prevalent infection and the vaccine efficacy on prevalent viral load. Pleasingly, these metrics should be better proxies for the effect of vaccine on transmission than traditional metrics which focus on the effect of a vaccine on individual infection and individual mean viral load irrespective of duration. For transmission, what matters is the transmission potential induced by the vaccine on any given day and the new metrics naturally capture this aspect. We recommend that future studies, whether cross-sectional or longitudinal sample, use these metrics to help describe the manifold effects of a vaccine.

In general, longitudinal sampling will be much more efficient than single point sampling as many more infections will be captured. In some studies, cross-sectional sampling will not be feasible as too few cases will be accrued. Nonetheless, for large studies where longitudinal sampling is not feasible and there is a high ‘capture’ rate of infections, cross-sectional sampling can have adequate power to test for non-null effects of VE_*PI*_ and VE_*PV L*_. For example with 90 infection cases from point sample(s) we have 90% power to detect a VE_*PI*_ of 0.50. Thus if longitudinal sampling for infection is not logistically feasible, cross-sectional sampling should be considered as a practical design that allows estimation of the effect of vaccine on proxies for transmission.

Future work could explore how to augment a cross-sectional study with subsequent daily sampling of infected volunteers, known as a prevalent cohort study Brookmeyer *and others* (1987), Degruttola *and others* (1991). Here serial viral loads and time to infection cessation would be collected. While our results appear robust to the constant attack rate assumption for COVID-19, for other diseases with varying attack rates and longer durations, this may be less plausible (e.g. HIV and AIDS). Work on how to weaken this assumption could also be explored. Finally, while our work focused on the binary outcome of infection, in some settings one can subdivide infections into those that result in disease versus those that are purely asymptomatic. Such subdivision could lead to a multinomial regression model for the two competing events of infection that progresses to diseases versus infection without disease.

## Data Availability

The data in the manuscript is available.

## Acknowledgments

We thank Elizabeth Brown, Peter Gilbert, Holly Janes, Jing Qin, and Stephanie Schrag for helpful comments.

## APPENDIX

Our development has been for the setting where the attack rate is assumed roughly constant for the period prior to the cross-sectional sample. The incidence of COVID-19 has changed over the course of the pandemic. To assess the the sensitivity of our simulation results to the constant attack assumption we did some simulations. Our duration distribution has a maximum duration of about 30 days. In the US over the course of the pandemic the largest change in incidence over any 30 day period has been about a tripling. We thus evaluate two scenarios. Under the increasing scenario, the attack rate at day 100 was 3 times the attack rate at day 70. Under the decreasing scenario, the attack rate at day 100 was 1/3 the attack rate at day 70. For each simulated study, we generate 70 infections over the period from day 70 to day 100 under the increasing or decreasing scenario and sampled PCR+ infections at day 100.

Table A1 below reports the results. We do see some bias in the sampled mean viral loads. Under the constant attack rate scenario of Table 1, and with peak VL and duration of 6 and 28, respectively, the mean mean sampled viral loads were about 3.0. With a peak VL of 4, the mean viral load was about 2.0 (regardless of duration). With an increasing attack rate these mean viral loads increase to about 3.4 and 2.3, respectively or an increase of about 15%. With a decreasing attack rate the mean viral loads decrease to about 2.7 and 1.8 or about at 10% decrease.

Under both scenarios, VE_*PI*_ appeared unbiased if the vaccine had no effect on duration. Under the increasing (decreasing) attack rate scenario VE_*PI*_ was slightly biased downward (upward) if the vaccine reduced the duration. The bias was less than 10%, often less than 5%. A similar pattern but with less bias was observed for VE_*P V L*_.

**Table A1.** Simulated performance of the simple cross-sectional estimates of VE_*PI*_ and VE_*PV L*_. The mean number of infections per arm is recorded along with the mean VEs and viral loads. Each row is based on 1,000 simulated trials. The placebo arm has a mean peak viral load of 6 and a mean duration of 28 days for all scenarios.

| Inputs |  |  |  |  | Performance |  |  |  |  |  |
| --- | --- | --- | --- | --- | --- | --- | --- | --- | --- | --- |
| VE<sub>I</sub> | VE<sub>PI</sub> | VE<sub>PVL</sub> | Peak | Infection | # Positive | | $\widehat{VE}_{PI}$ | $\widehat{VE}_{PVL}$ | $\overline{V}_0$ | $\overline{V}_1$ |
|  |  |  | VL | Duration | Vaccine | Placebo |  |  |  |  |
| Attack Rate at Day 100 is 3 times that of Day 70 |  |  |  |  |  |  |  |  |  |  |
| 0.00 | 0.00 | 0.00 | 6 | 28 | 33.30 | 33.20 | -0.033 | -0.044 | 3.42 | 3.42 |
| 0.50 | 0.75 | 0.75 | 6 | 14 | 13.50 | 44.40 | 0.689 | 0.702 | 3.41 | 3.25 |
| 0.50 | 0.50 | 0.67 | 4 | 28 | 22.30 | 44.40 | 0.485 | 0.651 | 3.41 | 2.29 |
| 0.50 | 0.75 | 0.83 | 4 | 14 | 13.50 | 44.40 | 0.689 | 0.800 | 3.41 | 2.18 |
| 0.75 | 0.88 | 0.88 | 6 | 14 | 8.01 | 53.20 | 0.847 | 0.855 | 3.42 | 3.20 |
| 0.75 | 0.75 | 0.83 | 4 | 28 | 13.30 | 53.20 | 0.744 | 0.829 | 3.42 | 2.28 |
| 0.75 | 0.88 | 0.92 | 4 | 14 | 8.01 | 53.20 | 0.847 | 0.903 | 3.42 | 2.14 |
| Attack Rate at Day 100 is 1/3 times that of Day 70 |  |  |  |  |  |  |  |  |  |  |
| 0.00 | 0.00 | 0.00 | 6 | 28 | 30.70 | 30.70 | -0.032 | -0.046 | 2.70 | 2.69 |
| 0.50 | 0.75 | 0.75 | 6 | 14 | 7.83 | 40.90 | 0.804 | 0.782 | 2.69 | 2.94 |
| 0.50 | 0.50 | 0.67 | 4 | 28 | 20.60 | 40.90 | 0.483 | 0.648 | 2.69 | 1.81 |
| 0.50 | 0.75 | 0.83 | 4 | 14 | 7.83 | 40.90 | 0.804 | 0.854 | 2.69 | 1.96 |
| 0.75 | 0.88 | 0.88 | 6 | 14 | 4.70 | 48.90 | 0.902 | 0.893 | 2.68 | 2.91 |
| 0.75 | 0.75 | 0.83 | 4 | 28 | 12.30 | 48.90 | 0.743 | 0.824 | 2.68 | 1.82 |
| 0.75 | 0.88 | 0.92 | 4 | 14 | 4.70 | 48.90 | 0.902 | 0.928 | 2.68 | 1.95 |

FREEMAN, JONATHAN AND HUTCHISON, GEORGE B. (1980). Prevalence, incidence and duration.

